# Mathematical Modelling the Impact Evaluation of Lockdown on Infection Dynamics of COVID-19 in Italy

**DOI:** 10.1101/2020.07.27.20162537

**Authors:** S. Kadyrov, A. Orynbassar, H.B Saydaliyev, D. Yergesh

**Affiliations:** Suleyman Demirel University, Kaskelen, Kazakhstan

**Keywords:** COVID-19, Lockdown, Epidemic model, SEIR, Awareness, Dynamical systems

## Abstract

The Severe Acute Respiratory Syndrome Coronavirus 2 (SARS-CoV-2), the cause of the coronavirus disease-2019 (COVID-19), within months of emergence from Wuhan, China, has rapidly spread, exacting a devastating human toll across around the world reaching the pandemic stage at the the beginning of March 2020. Thus, COVID-19’s daily increasing cases and deaths have led to worldwide lockdown, quarantine and some restrictions. Covid-19 epidemic in Italy started as a small wave of 2 infected cases on January 31. It was followed by a bigger wave mainly from local transmissions reported in 6387 cases on March 8. It caused the government to impose a lockdown on 8 March to the whole country as a way to suppress the pandemic. This study aims to evaluate the impact of the lockdown and awareness dynamics on infection in Italy over the period of January 31 to July 17 and how the impact varies across different lockdown scenarios in both periods before and after implementation of the lockdown policy. The findings SEIR reveal that implementation lockdown has minimised the social distancing flattening the curve. The infections associated with COVID-19 decreases with quarantine initially then easing lockdown will not cause further increasing transmission until a certain period which is explained by public high awareness. Completely removing lockdown may lead to sharp transmission second wave. Policy implementation and limitation of the study were evaluated at the end of the paper.

## 1 Introduction

In late December 2019 a novel coronavirus (COVID-19) pneumonia outbreak occurred first time in Wuhan, Hubei Province, China which shortly became pandemic affecting entire world. Being zoonotic coronavirus, COVID-19 is found to be severe respiratory illness similar to Middle East respiratory syndrome MERS-CoV from 2012 and severe acute respiratory syndrome SARS-CoV from 2002. The possible occurrence of novel coronavirus epidemic in China was predicted about a year ago in a research review [3] published on 2 March 2019.

As of July 23, 2020 there are over 15.4 million total confirmed cases in the world, over 631 thousand deaths, and about 9.4 million recovered. Italy is one of the countries that had rapid growth of the confirmed cases in the early days since the first case in the country. In January 31, the first COVID-19 cases were detected in Italy when two Chinese tourists tested positive. As of July 23, 2020 the total laboratory-confirmed COVID-19 cases is reported to be 245,032, with death toll raising to 35,082, and 197,628 recovered cases. See Figure 1 for the progression of the virus in Italy.

**Fig. 1:**
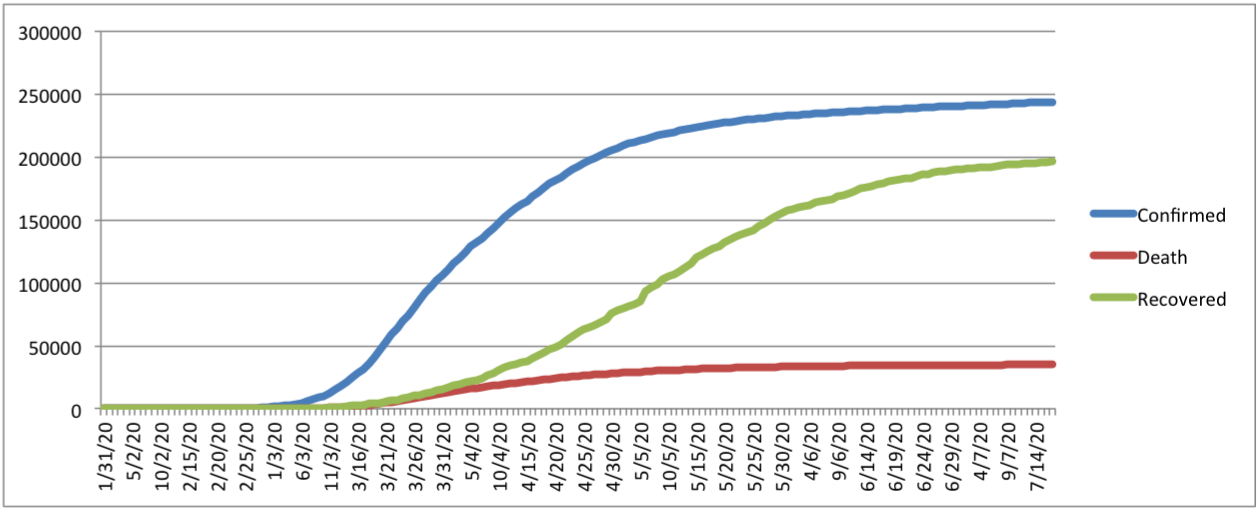
Italy COVID-19 data from January 31, 2020 to July 17, 2020

In an unprecedented effort to contain the pandemic, the nations of the world have implemented strict measures at the core of which is social distancing and travel bans. The efficacy of these measures depends, among other factors, on the capacity for rapid identification of cases and tracking the rapidly evolving SARS-CoV-2 lineages. As the search for vaccine and effective treatments continues, early stratification of COVID-19 patients by disease severity may aid more effective allocation of healthcare resources and reduce undesirable clinical outcomes. This could be facilitated by the application of COVID-19-associated biomarkers predictive of disease outcomes.

After 38 days since the first confirmed case, on March 8, the Lombardy region in Italy, along with 14 other provinces in the north and center of Piedmont, Emilia-Romagna, Veneto and Marche, were put under lockdown. Two days later, the government extended blocking measures to the whole country. Two weeks later, the number of new cases per day showed signs of slowing, while the number of new deaths increased slightly.

On March 31, the president of the National Institute of Health, Silvio Brusaferro, announced that the pandemic in the country had reached its peak. The news was also confirmed by the chief of Civil Protection, Angelo Borrelli.

Three weeks after the lockdown, its consequences became apparent. Italy reported a decrease in new cases and new deaths per day. The country is also experiencing a constant reduction in intensive care unit personnel. On April 5, Italy had the smallest number of new daily deaths in two and a half weeks, and every other day the smallest number of new daily deaths in three weeks. On April 20, 2020, Italy registered the first decline in the number of active cases.

Starting from May 18, the Italian government started easing the lockdown restrictions allowing most businesses to reopen and letting free movement to all citizens.

In this study we consider a version of a deterministic SEIR epidemic model to estimate epidemiological parameters of COVID-19 in Italy when lockdown measures and awareness against the pandemic are taken into account. We then use these parameters to predict how the situation progresses under different scenarios and whether the second wave will occur when the lockdown is removed.

## 2 Methodology

### 2.1 Data

We use dataset from January 31, 2020 when first positive cases reported in Italy till July 17, 2020. To collect Italy COVID-19 time series data of confirmed, recovered, and death cases we use COVID-19 Data Repository by the Center for Systems Science and Engineering (CSSE) at Johns Hopkins University [5].

### 2.2 Model

We split the whole population into four compartments: Susceptible (*S*), Exposed (*E*), Infectious (*I*), and Recovered (*R*). We consider a version of SEIR epidemic model with model diagram given in Figure 2.

**Fig. 2:**
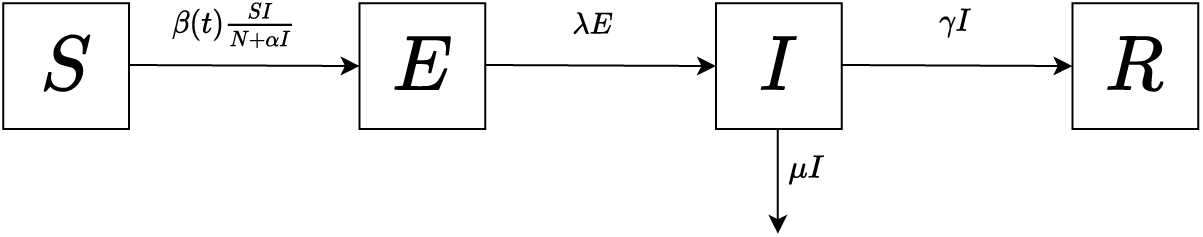
SEIR model diagram

A corresponding system of ordinary differential equations it takes the form

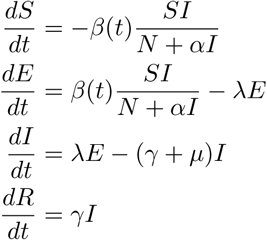

where *N* = 60480000 is the total population of Italy, *β*(*t*) is the time dependent transmission rate, *λ* the daily rate of exposed individuals becoming infectious, *µ* the rate of death due to virus, *γ* the recovery rate from virus, and *α* can be thought of as parameter to measure the awareness of the population against the pandemic. As noted above, all parameter except *β*(*t*) are assumed to be constant. The reason for time dependence of *β*(*t*) is mainly due to quarantine and lockdown measures that we aim to take into account. To be precise, we assume an exponential lockdown function between [*t*_0_, *t*_1_] time intervals with an exponent 𝓁> 0.

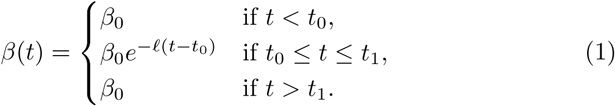

See Figure 3 for the graph of our lockdown function. Here we are assuming a uniform lockdown, while some studies consider different kind of lockdown, see e.g. [4] where some individuals are separated from *S* into protected compartment.

**Fig. 3:**
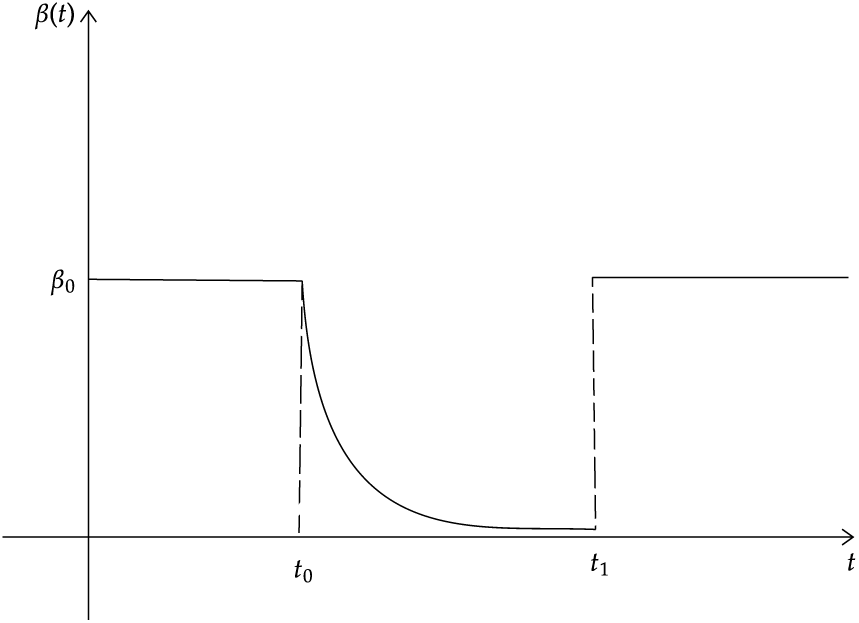
Lockdown function

### 2.3 Estimation

We note that the parameter *λ* is taken to be the inverse of incubation period. Recent studies estimate the mean incubation period to be 6.4 days (95% CI, 5.6 to 7.7) [1] and median incubation period to be 5.1 days (95% CI, 4.5 to 5.8) [7]. For our purposes we assume that *λ* = 1*/*6.4.

While early reports of Chinese Center for Disease Control and Prevention [2] suggests the infectious period to be 9 days, another recent work [11] reports the mean infectious period to be 10.91 days (SD=3.95). For our purposes we take the parameter *γ* = 1*/*10.91.

To estimate the daily death rate *µ* we use simple linear regression. To this end, let *D*(*t*) denote the total number of deaths up to date *t*. Then, we can model the daily rate of change with *dD/dt* = *µI*, which can be approximated by

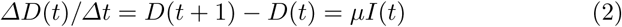

as Δ*t* = 1.

Finally the remaining parameters *β*(*t*), *α* are estimated from nonlinear least square optimization technique. More specifically, we use ‘scipy.integrate.ode function’ of python programming language to simulate S(t), I(t), E(t), and R(t) where we let initial values as S(0)=country population, I(0)=the first observed number of cases in the country, E(0)=0, and R(0)=0. Then, we ‘call scipy.optimize.curve_fit’ function for least-square fitting of the theoretical model solution to the observed daily number of confirmed cases and recovered individuals.

## 3 Results

### 3.1 Death rate *µ*

To compute death rate we compute Δ*D* from the data and use the linear equation (2). Since *µ* is assumed to be constant, we want to use the linear regression analysis. As seen from scatter plot in Figure 4a the graph is not linear. However, if we restrict to the first 50 days then we see the linear relation Figure 4b.

**Fig. 4:**
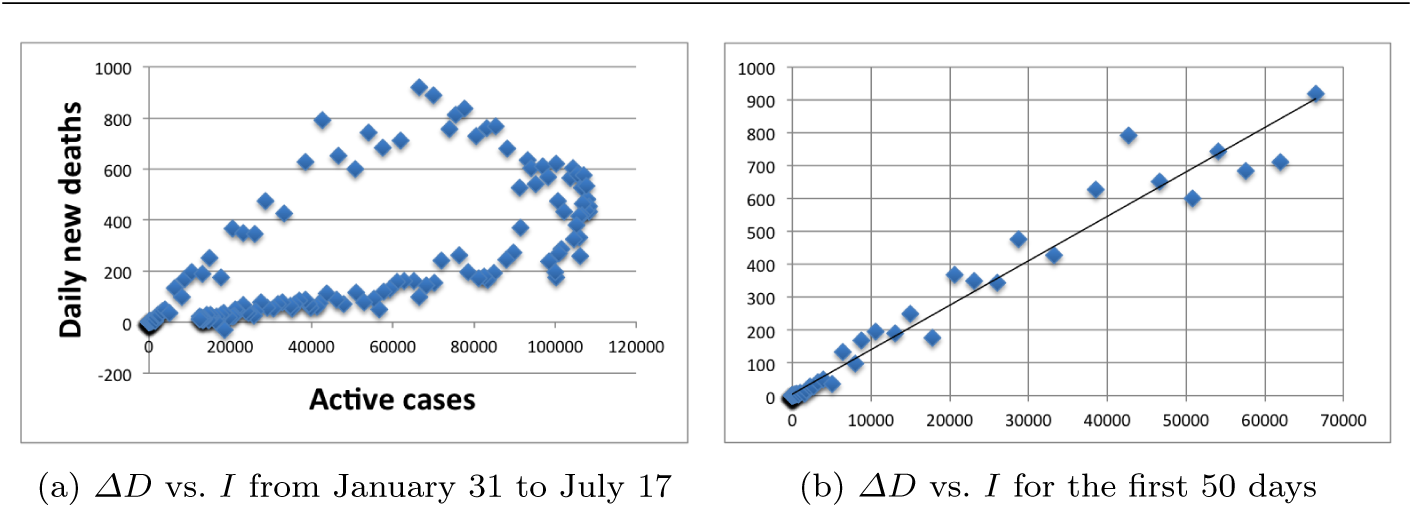
A figure with two subfigures

Applying simple linear regression with zero intercept to the first 50 days we obtain *µ* = 0.01365 (95% CI, 0.01308 to 0.01423). This is what we use for *µ* to estimate the remaining parameters.

### 3.2 Transmission rate *β* without lockdown

We now estimate *β*(*t*) given in (1). To this end, we first estimate the constant coefficient *β*_0_ by restricting ourselves to the first 38 days (January 31-March 8) when there were no lockdown measures applied. So, we may safely assume *β*(*t*) = *β*_0_ on [0, 38] and also set awareness *α* = 0. Figure 5 shows the plot of data points and estimated curve for the active cases. Parameter estimation from the SEIR model using the least squares gives *β* = 0.8546, (95% CI, 0.8444 to 0.8648).

**Fig. 5:**
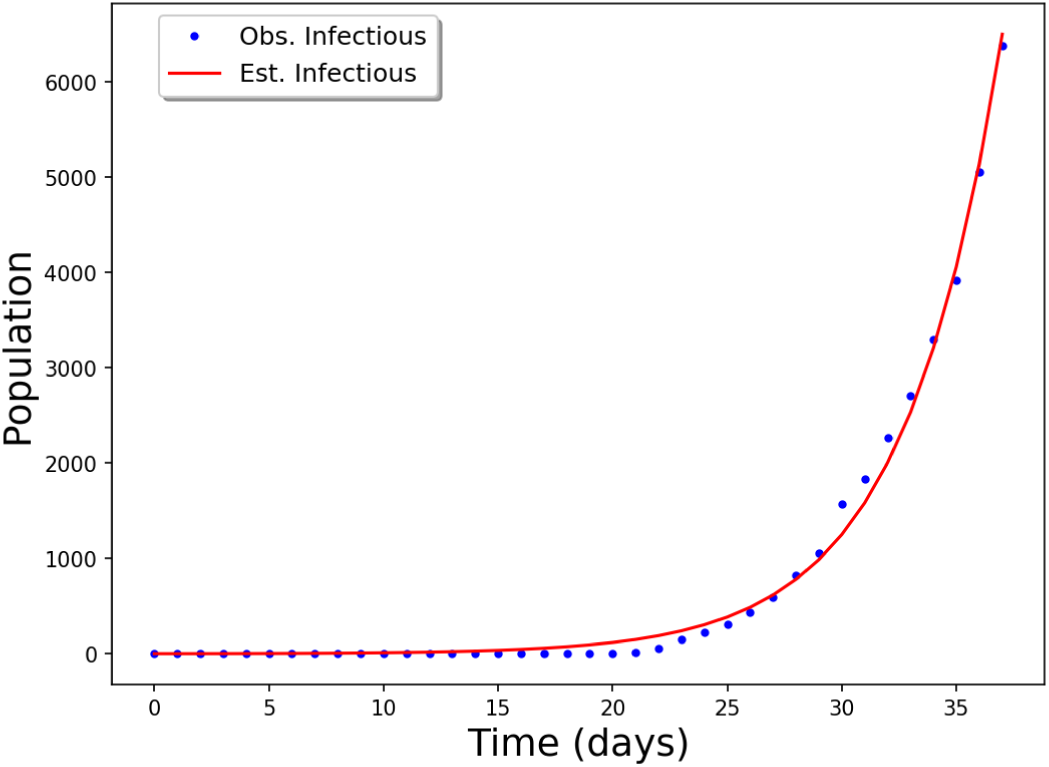
Plot of the observed and simulated active cases for the first 38 days

### 3.3 Transmission rate *β* with lockdown and awareness estimates

Now that we have *β*_0_ estimated, we need to estimate the remaining parameters of *β*(*t*) from (1), namely, *t*_0_, the start of lock down, 𝓁, the exponent. To do so, we use the dataset from January 31 to May 18, when the lockdown measures eased. In particular, we safely let *t*_1_, the end of lockdown larger that May 18. The estimated active cases shown in Figure 6. The point estimates are 𝓁 = 0.0243, (95% CI, 0.0219 to 0.0267), *t*_1_ = 67.5, (95% CI, 65.4 to 69.6), and *α* = 3230.4, (95% CI, 3112.0 to 3348.8).

**Fig. 6:**
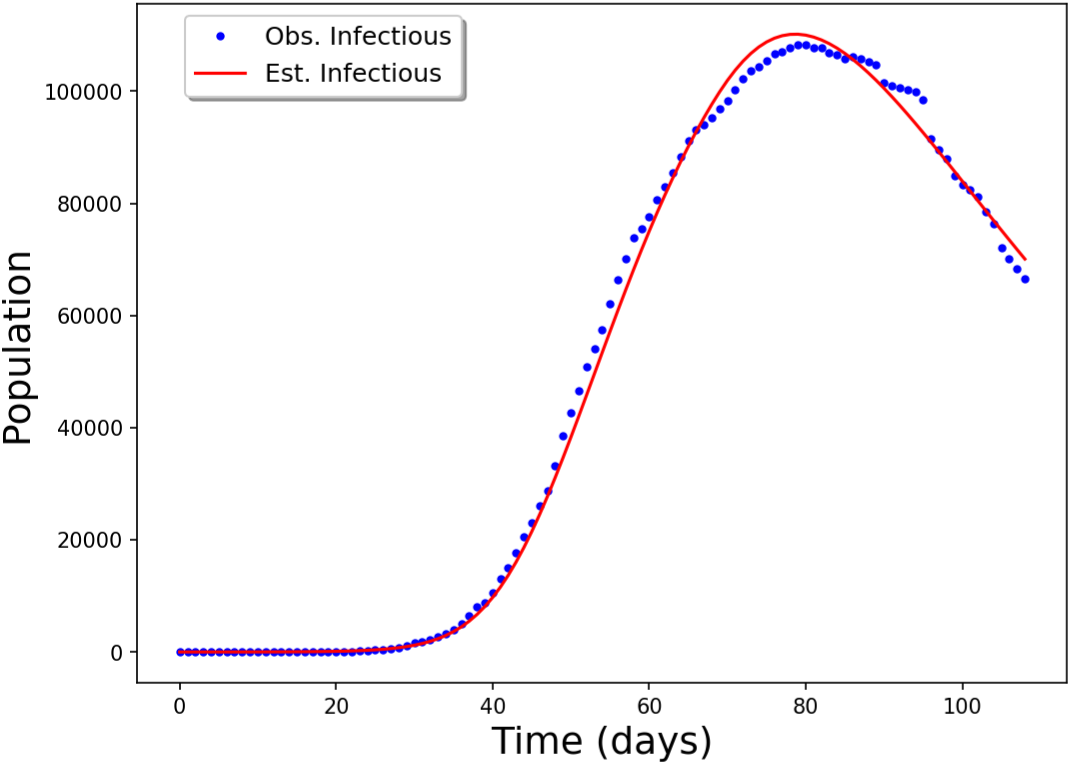
Plot of the observed and simulated active cases until the end of lockdown period

### 3.4 Forecasting possible scenarios after first lockdown

Using all the estimated parameters, we test the model to the dataset until July 17. Here we still assume that lockdown continues, that is, *t*_1_ is large in (1). Both estimated and observed active cases are shown in Figure 7. We see that the effect of the lockdown still continued until day 162 (July 10) and in the last week our model under estimated the observed active cases.

**Fig. 7:**
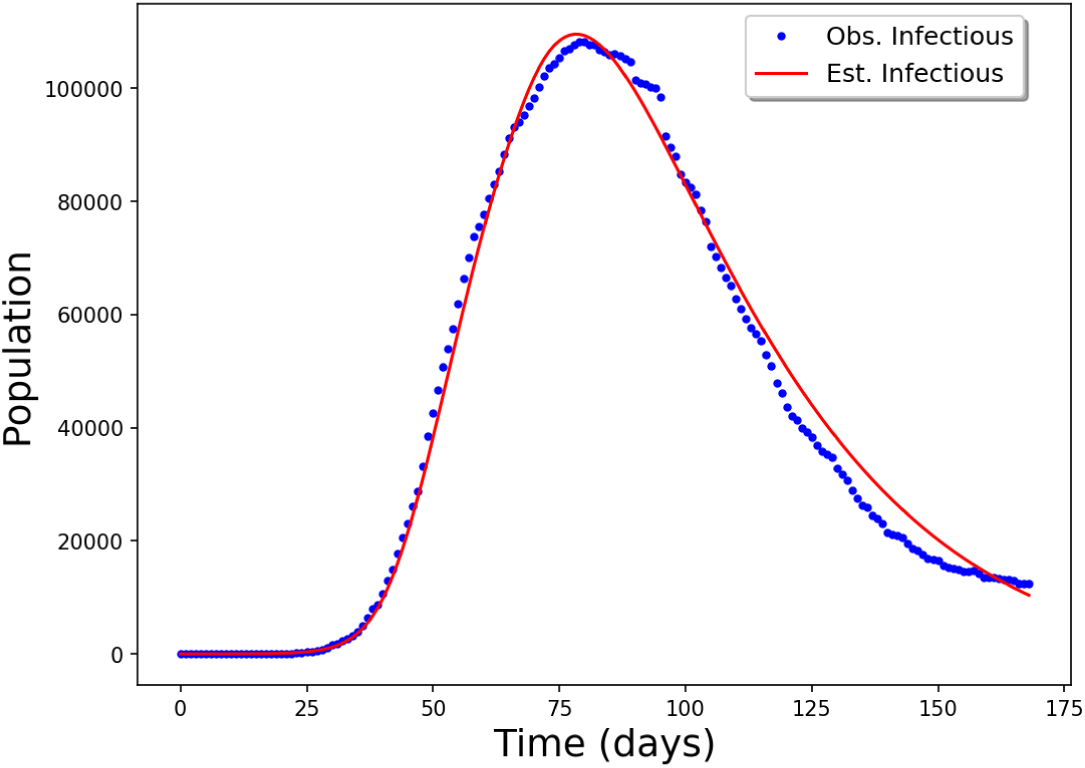
Plot of the observed and simulated active cases until July 17, 2020

If we completely remove the lockdown on July 10, then we see that the second wave is unavoidable, see Figure 8 with *ϵ* = 1. In fact, the second wave still occurs if the lockdown is removed completely in a later date. So, the better option would be to ease the lockdown but do not remove it completely. To show the effect of various lockdown scenarios we consider the following modified version of our lockdown function *β*:

**Fig. 8:**
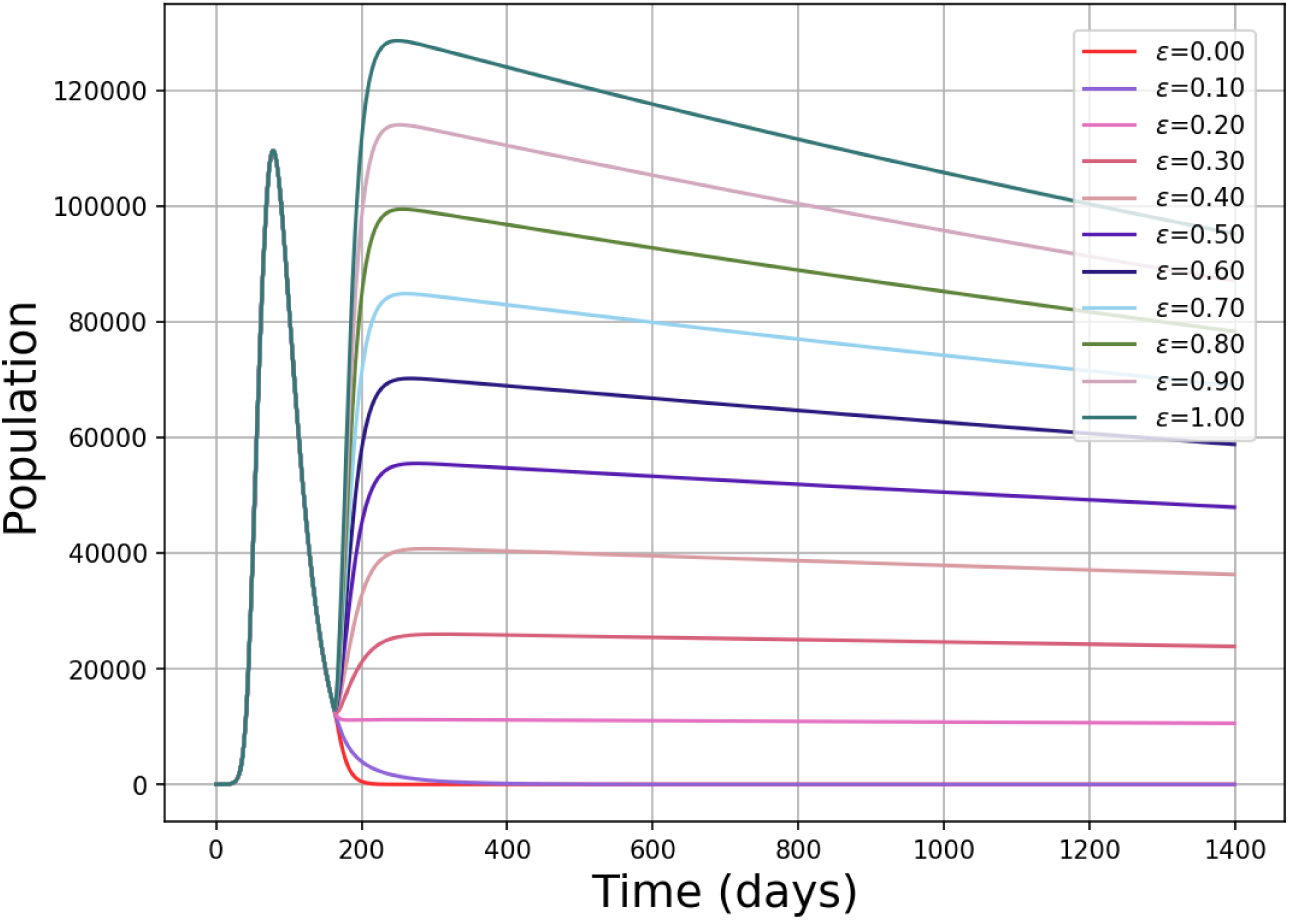
Possible lockdown scenarios for the future

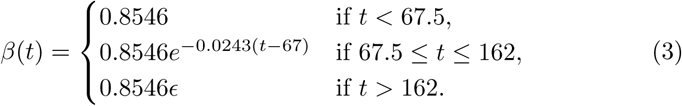

Here, the parameter *ϵ ∈* [0, 1] indicates the lockdown percentage after July 10, with *ϵ* = 0 meaning the complete lockdown and *ϵ* = 1 meaning no lockdown at all. For *ϵ* = 0.0, 0.1, 0.2, …, 1.0 we have Figure 8.

## 4 Discussion

In this paper we considered a modified SEIR model with lockdown and awareness taken into account to estimate COVID-19 epidemiological characteristics using Johns Hopkins University dataset for Italy over the period from January 31 to July 17. The summary of parameter estimation reveals that the death rate *µ* = 0.01365, transmission rate *β*_0_ = 0.8648 giving the basic reproduction number *R*_0_ = *β*_0_*/*(*γ* + *µ*) = 5.03. One of the limitations of the study was to estimate the death rate as a constant parameter using only first 50 days of the dataset. However, Figure 4a suggests the death rate is not a constant number but piecewise constant. It could be an interesting research problem to find out why there is a negative slope in the scatter plot.

As can be readily seen from Figure 6 and Figure 7 our lockdown function *β*(*t*) in (1) nicely approximated the observed active cases with exponent 𝓁 = 0.0243. In Italy, the start of imposing the lockdown measures started on March 8 which corresponds to *t* = 38 day since the first confirmed case. However, our estimates for the start of lockdown is *t*_1_ = 67.5. It maybe interpreted as the real effect of lockdown started appearing after 29 days since the start of the lockdown. This is consistent with the early reports that state that consequences of lockdown were apparent after three weeks.

Our analysis reveal that the awareness parameter is *α* = 3230.4. There is no upper bound for *α* and hence it is not clear how it should be interpreted, other than noting that awareness is high. We interpret *αI* term as awareness with increase of infectious the term increases and hence the incidence rate *β*(*t*)*SI/*(*N* + *αI*) decreases. On the other hand, some studies interpret it as “the measure of inhibition” taken by infectious individuals [6].

Based on the results and findings of this study, a few policy implications were determined. Firstly, with respect to the objective of this study, implementation of the lockdown has minimized the social distancing flattening the curve. We see that Italy should not remove the lockdown completely. The policy implication of this finding involves the need to impose lockdown policy to some extend otherwise any relaxation in the lockdown may lead to sharp transmission second wave.

Each model has some assumptions, some very much depends on the public’s compliance to the regulations set by the government. The model based on dynamical systems can incorporate more parameters in its simulation such as contact rate and duration of recovery to help understand the pattern of spread. The curve-fitting model, despite its reported accuracy, is too dependent on the historical data. The true reflection of reality can only be achieved if more tests can be performed. The SEIR model is simple and only considers 4 parameters, which are all inferred from the historical data.

## 5 Conclusion

The COVID-19 epidemic was reported in the Hubei province in China in December 2019 and then spread around the world reaching the pandemic stage at the beginning of March 2020. Since then, several countries went into lockdown. COVID-19 epidemic in Italy started as a small wave of 2 cases in January 31 through imported cases. It was followed by a bigger wave mainly from local transmissions resulting in 6387 cases. The following wave saw unexpectedly three digit number of daily cases following a mass gathering urged the government to choose a more stringent measure. A lock-down approach was immediately initiated at 8 March to the whole country as a way to suppress the epidemic trajectory. The lock-down causes a major socio-economic disruption thus the ability to forecast the infection dynamic is urgently required to assist the government on timely decisions. Limited testing capacity and limited epidemiological data complicate the understanding of the future infection dynamic of the COVID-19 epidemic. The objective of this study was to evaluate the impact of the lockdown dynamics on infection in Italy and how the effects vary across different lockdown scenarios. This study attempted to achieve objective using the SEIR estimation technique based on a Johns Hopkins University dataset for Italy from January 31 to July 17. We tried to measure for both periods before and after the implementation of the lockdown policy. The obtained results of the comparison study showed that implementation of the lockdown has been minimized due to the social distancing can flatten the curve.

## Data Availability

we use COVID-19 Data Repository by the
Center for Systems Science and Engineering (CSSE) at Johns Hopkins University

## Acknowledgements

The first author acknowledges the grant from the Ministry of Education and Science of the Republic of Kazakhstan within the framework of the Project AP08051987.

